# Cost-Effectiveness Analysis of Deep Brain Stimulation for the Treatment of Alcohol Use Disorder and Alcoholic Liver Disease

**DOI:** 10.1101/2024.08.22.24312455

**Authors:** O.A. Abiola, C. Lehmann, K. Moussawi, H. Jalal

## Abstract

**Background:** Alcohol use disorder (AUD) is a major public health concern and cause of mortality and morbidity. Alcohol-associated liver disease (ALD) is a debilitating complication of AUD, mitigated by abstinence from alcohol use. Deep brain stimulation (DBS) is emerging as a potential treatment for AUD. However, its cost-effectiveness compared to the standard medical treatment is unclear.

**Objective:** To estimate the cost-effectiveness of DBS compared to medical management for patients with AUD and ALD.

**Methods:** We utilized a decision analytic model based on published literature to conduct a cost-effectiveness analysis of costs and health outcomes for DBS and medical management in patients with AUD and ALD. We also carried out a threshold analysis to determine the probability of success necessary for DBS to be cost-effective. Costs were measured in 2024 US dollars and effectiveness in quality-adjusted life years (QALYs). We used a time horizon of 1-2 years and adopted a societal perspective.

**Results:** Our results show that for AUD patients in general, DBS is not cost-effective at any DBS success rate. However, for advanced ALD patients, defined as fibrosis stage 3 or beyond DBS becomes cost-effective. For these patients, DBS is cost-effective over a two-year period at a $100,000 willingness-to-pay threshold at DBS success rates greater than 53%. For advanced decompensated ALD patients, DBS is cost-effective over a one-year period at DBS success rate greater than 35%.

**Conclusion:** Should it prove efficacious, DBS may be cost-effective for patients with AUD and ALD. Thus, future randomized controlled trials to evaluate its efficacy are warranted.

## Introduction

Alcohol is the most misused substance globally and is a major public health problem [1]. More than 10% of the US population older than 12 years old have alcohol use disorder (AUD). Each year, the economic burden from alcohol misuse is estimated at $250 billion and more than 95,000 otherwise preventable deaths are related to AUD [2]. A quarter of these deaths are due to advanced alcohol-associated liver disease (ALD) [3–6]. Importantly, the prevalence and severity of ALD and other AUD-associated health problems increased since the COVID-19 pandemic [7–9].

Excessive and chronic alcohol consumption (> 40g/day) invariably results in ALD [6, 10–12] and continued alcohol consumption leads to more severe stages of ALD. As ALD develops, the liver histopathology progresses through four stages of fibrosis starting with alcoholic fatty liver (stage 1) to alcoholic cirrhosis (stage 4). Nearly 15% of all AUD patients develop alcoholic cirrhosis in their lifetime, which can be compensated (asymptomatic) or decompensated (symptomatic). Cirrhosis is associated with an overall mortality risk of 50% within 5 years [6, 11, 13–18]. Importantly, stage 3 fibrosis marks a turning point in terms of clinical prognosis as unlike early stage ALD (fibrosis stages 1-2), advanced ALD (fibrosis stage ≥ 3) is associated with 10-year mortality of 45% when compensated, and 93% mortality when decompensated [19, 20].

Abstinence from alcohol use improves ALD and reduces mortality with permanent abstinence being the ultimate treatment for all stages of ALD. Abstinence reverses histological features of ALD, improves clinical outcomes, and lowers mortality rates, independent of the clinical stage of ALD [6, 13, 15, 16]. For example, in patients with alcoholic cirrhosis, abstinence improves the 5-year survival from about 50% up to 87% [18, 21].

However, the rate of abstinence in AUD remains very low even with available treatments. FDA-approved treatments for AUD include disulfiram, acamprosate, and naltrexone, which are often used in combination with behavioral therapy [22]. Despite these interventions, the overall relapse rate to alcohol use has not improved over the past 50 years (55% relapse rate within 6 months) [23, 24]. Further, 90% of patients with AUD relapse within a year [25]. Hence, identifying novel and more efficacious treatments to prevent relapse to alcohol use is an urgent public health priority.

Deep Brain Stimulation (DBS) is emerging as a potential treatment for substance use disorders including AUD [26–28]. More than 208,000 DBS systems have been implanted so far, mostly for fully FDA-approved indications like Parkinson’s disease (PD), epilepsy, and essential tremor, and for other indications under a Humanitarian Device Exemption (e.g., dystonia, obsessive compulsive disorder (OCD), Tourette’s syndrome, Lesch-Nyhan syndrome) [29–33]. DBS involves the implantation of stimulating electrodes in the brain, which are controlled by a neurostimulator, and is thus an expensive and higher-risk intervention than currently available treatments for AUD. Studies on the efficacy of DBS for AUD remain preliminary and the success rate of DBS as a treatment for AUD has not been established. As such, this study aims to evaluate the cost-effectiveness of DBS for the treatment of AUD (general population) and AUD patients with ALD. In addition, given the increased risk associated with DBS surgery in these patients, we conduct a threshold analysis to determine at which DBS success rates, DBS becomes cost-effective when accounting for increased DBS-related risk.

## Materials and Methods

### Study Design

We constructed a decision tree to compare the costs and effectiveness of standard medical management (MM) with a hypothetical DBS treatment (Fig. 1). The base-case scenario consisted of 3 AUD patient-groups with or without ALD choosing to undergo treatment with either MM or DBS: 1) patients with AUD, 2) patients with advanced ALD (> moderate fibrosis), and 3) patients with advanced decompensated ALD (i.e., symptomatic cirrhosis). A patient who chooses to undergo MM is enrolled in a 16-week-long treatment that entails a combination of FDA-approved drugs, naltrexone and acamprosate, in addition to behavioral therapy. This combination was chosen as it was shown to be more efficacious than monotherapy [34]. A patient who chooses DBS undergoes an invasive neurosurgical intervention that targets the nucleus accumbens [26, 27]. We estimated the costs and utilities of MM and DBS under different probabilities of treatment success.

**Fig. 1.**
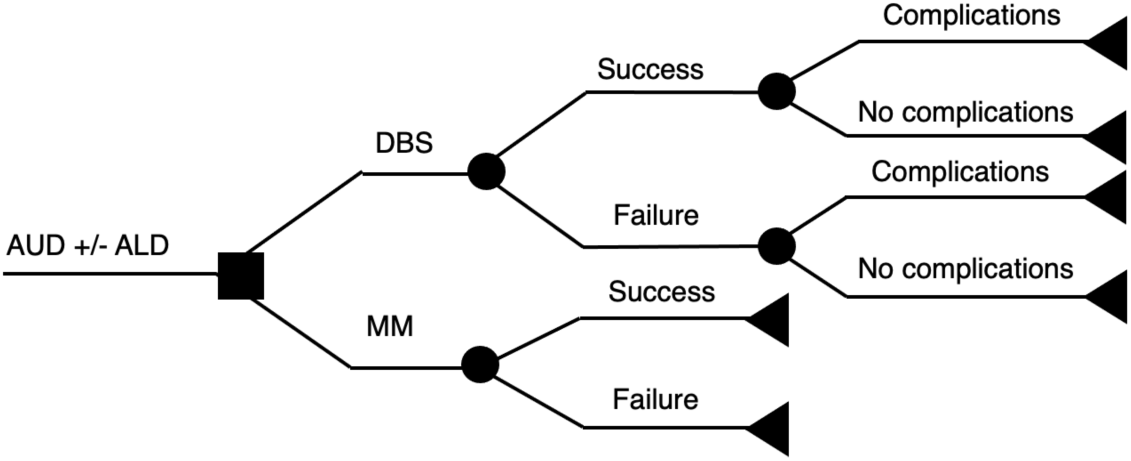
Model Decision Tree

Both interventions result in one of two outcomes: success or failure. Success here is defined as complete abstinence or moderate drinking without problems a year after the intervention, where moderate is defined as a maximum of 11 drinks (for women) or 14 drinks (for men) per week, with no more than 2 days on which more than 3 drinks (for women) or 4 drinks (for men) were consumed. Conversely, failure is defined as completing treatment but not abstaining or having more than moderate amounts of alcohol. These outcomes were defined by the Combined Pharmacotherapies and Behavioral Interventions for Alcohol Dependence (COMBINE) study [35] – the largest current alcohol dependency study – and were adapted to the DBS arm to show consistency.

The proposed model accounts for common complications associated with DBS surgery. However, given the increased surgical risk in patients with AUD and ALD, we carried out additional analyses to study the cost-effectiveness of DBS with higher complication risks than those commonly associated with DBS surgery [36, 37].

### Model Inputs (Table 1)

#### Probabilities

For MM, probabilities of DBS success were calculated based on the outcomes of the COMBINE study [35]. The study involved AUD patients (n = 1383) who were assigned to one of nine MM combinations and compared the effectiveness of each on three metrics: percent days abstinent (PDA), good clinical outcome, and relapse to heavy drinking. For our analysis, we used the relapse to heavy drinking outcome as the success/failure threshold. The COMBINE study followed up with patients a year after the treatment, allowing the estimation of success probability of MM (Table 1). Of note, the COMBINE study excluded AUD patients with any ALD.

**Table 1.** Probabilities, QoL, and costs associated with the treatment of alcohol use disorder and alcoholic liver disease.

| Variable | Mean | Reference |
| --- | --- | --- |
| Probability of MM leading to abstinence | 0.246 | Anton et al. [35] |
| QoL at baseline |  |  |
| AUD | 0.49 | Laaksonen et al. [39] |
| Advanced ALD | 0.43 | Janani et al [40] |
| Decompensated ALD | 0.40 | Janani et al [40] |
| QoL with abstinence |  |  |
| AUD | 0.66 | Laaksonen et al. [39] |
| Advanced ALD | 0.57 | Estimated |
| Decompensated ALD | 0.52 | Estimated |
| Cost of MM | \$1,530 | Zarkin et al. [34] |

#### Quality of Life

To determine the effectiveness of each treatment a year from baseline, we compared the quality-adjusted life years (QALY) gained by patients one year after the intervention. Quality-of-life (QoL) studies and estimates most commonly report values over one year, thus we chose to construct the majority of our analyses over this time horizon. In addition, we carried out a 2-year analysis for patients with AUD and advanced ALD to estimate QALYs gained 2 years after the intervention for that patient population. QALYs are calculated by multiplying the QoL of a patient and the duration of time they have that QoL [38]. To quantify the utility directly lost or gained due to treatment failure or success, we adopted values from a clinical trial that recorded the QoL of patients who had AUD before their intervention and after they no longer met the diagnosis requirement (Table 1) [39]. For ALD patients, given that there are very few prevalence-based measures of QoL published, we had to rely on estimates and non-alcoholic liver disease values [40]. The estimated QoL values with abstinence for ALD patients were calculated by increasing the baseline QoL values by the same percent increase (34.67%) found in AUD patients. All outcome QoL values accounted for the 2.63% standard risk associated with DBS surgery, which was estimated based on incidence and complication rates of DBS in movement disorders studies [41].

#### Costs

MM costs were highlighted in the COMBINE study [35] and adjusted to 2024 US dollars for our analysis (Table 1). Each hypothetical MM patient was assigned one of two costs: costs of treatment alone for those who had a successful intervention and (societal + treatment) costs for those with failed interventions. Similar methods were adopted for DBS patients, substituting costs and complications of MM with those of DBS (Table 2) [42]. Physician payment, hospital stay, and associated complications were all accounted for.

**Table 2.** Costs associated with DBS procedure adapted to 2024 US dollars. Adopted from Kuijper et al.

| Category | Cost (\$) |
| --- | --- |
| Diagnostic imaging and planning |  |
| CT | \$53.03 |
| MRI | \$92.91 |
| Lead Implantation |  |
| With microelectrode recording, one array | \$2,922.61 |
| Extra array | \$645.53 |
| Generator implantation |  |
| Connection to two or more arrays | \$1,077.62 |
| Analysis and programming (one year) | \$353.22 |
| Physician price for two electrodes | \$5,964.87 |
| Implantation: whole system, multiple array |  |
| No complication | \$28,115.80 |
| CC or MCC | \$40,322.97 |
| Total price for two electrode without CC/MCC | \$34,080.65 |
| Total price for two electrode with CC/MCC | \$46,287.84 |

Direct and indirect costs associated with each condition were adapted from national reports and studies containing the direct healthcare costs, and indirect costs like lost productivity, non-healthcare consumption, and crime-related expenses [2, 43–45]. For AUD patients, the annual cost per individual was obtained by dividing the total societal cost by the number of people with AUD in 2010 and adjusting to 2024 US dollars using the Bureau of Labor Statistics Consumer Price Index inflation calculator (Table 3) [2].

**Table 3.** Societal costs associated with different patient populations adapted to 2024 US dollars. Adapted from Rehm et al and Julien et al.

| Category | Cost (\$) |
| --- | --- |
| Healthcare Costs |  |
| AUD | \$2,238.29 |
| Advanced ALD | \$30,198.48 |
| Decompensated ALD | \$135,172.58 |
| Lost Productivity Costs |  |
| AUD | \$15,101.59 |
| Advanced ALD | \$60,396.96 |
| Decompensated ALD | \$270,345.17 |
| Total Costs |  |
| AUD | \$22,676.15 |
| Advanced ALD | \$90,595.44 |
| Decompensated ALD | \$405,517.75 |

For both the advanced ALD and decompensated ALD groups, we approximated costs based a study that estimated total costs associated with various stages of ALD in 2022 and adjusted them to 2024 US dollars (Table 3) [44, 45]. Stages F3 and F4 were reported to be prevalent in 756,677 and 167,302 people, respectively, and had healthcare costs of $1,828 per person in the first year. With decompensated liver disease, the prevalence is lower with 90,933 people with ascites, 60,700 people with variceal bleeding, and 24,512 people with both. These decompensated ALD states have healthcare costs of $104,507, $108,819, and $213,325 per person respectively. Decompensated encephalopathy had a prevalence of 28,129 and costs $146,680. These costs were consistent previous reports [43].

#### Threshold and cost-effectiveness Analysis

We first performed a threshold analysis which was developed to determine the probability of success necessary for DBS to impart the same cost and effectiveness (measured in QALYs) as MM. This provided a baseline measure to compare the incremental cost-effectiveness ratios (ICER) of the difference in expected costs and expected QALYs [19]. These ICERs were compared to a range of willingness-to-pay thresholds ranging from $0 to $1 million per QALY such that for an intervention to be defined as cost-effective, the ICER must be lower than the willingness-to-pay. We present this data on a cost-effectiveness acceptability curve which shows the probability of each strategy being cost-effectiveness at various willingness-to-pay thresholds. Given that the probability of DBS success is unknown, we also plotted the data on a heatmap to account for the effect of both DBS success probability and willingness-to-pay thresholds on DBS cost-effectiveness.

A probabilistic analysis was conducted by running 10,000 Monte Carlo simulations to examine the effects of variability in the model inputs on our cost-effectiveness findings. Gamma distributions were used for costs and beta distributions for probabilities and QoL values. Our analyses were all carried out using Excel and MATLAB, and our study design and reporting are in line with what is recommended by the Consolidated Health Economic Evaluation Reporting Standards [46].

#### Risks and Complications

The standard risk of DBS has been estimated to be 2.63% based on movement disorders studies [41]. Thus, when exploring the effects of potentially increased surgical risk associated with AUD and ALD on the cost-effectiveness of DBS, we multiplied the expected percent decrease in QoL by up to 32 folds. We also accounted for the increased expected cost of DBS due to complications by including the costs of the QALYs lost. Given that a statistical human life is expected to cost $7.2 million, and the life expectancy of a healthy person in the US is 79 years, the cost of a perfect year of life is about $91,000 [47]. Thus, the increased potential risk associated with DBS resulted in a decrease in QoL and a corresponding increase in cost, both of which were accounted for.

## Results

### Expected QoLs and Costs

Given that 24.6% of AUD patients completed the 16-week MM intervention and were still defined as successful one year after baseline, the expected QoL values are 0.532 for AUD, 0.469 for advanced ALD, and 0.441 for decompensated ALD patients a year after the intervention. These values were calculated as the weighted average based on the above success rate with MM treatment course (e.g., (24.6 x 0.66 + 75.4 x 0.49)/100) (Table 1). While the intervention costs were consistent amongst all 3 patient populations (Table 1 and 2), the healthcare and societal costs differed substantially (Table 3).

### Sensitivity Analyses

We found that DBS imparts an equivalent increase in QoL as MM at success rates of 29.8%, 28.1%, and 27.9% for patients with AUD, advanced ALD, and decompensated ALD, respectively (Fig. 2). For AUD patients, at 29.8% DBS success rate, DBS-group costs were $86,000 compared to $17,280 for MM group. For advanced ALD, at 28.1% DBS success rate, DBS-group costs were $130,990 vs. $64,090 for MM group. Lastly, for decompensated ALD, at 27.9% DBS success rate, DBS-group costs were $339,660 vs. $282,130 for MM group (Fig 2.). At 100% DBS success rate, DBS-group costs were $71,310 for all patient populations. Given the small difference in the costs of DBS and MM at a 100% DBS success for the advanced ALD group, we carried out a two-year analysis and found that DBS is cost saving at higher success rates (>53%).

**Fig. 2.**
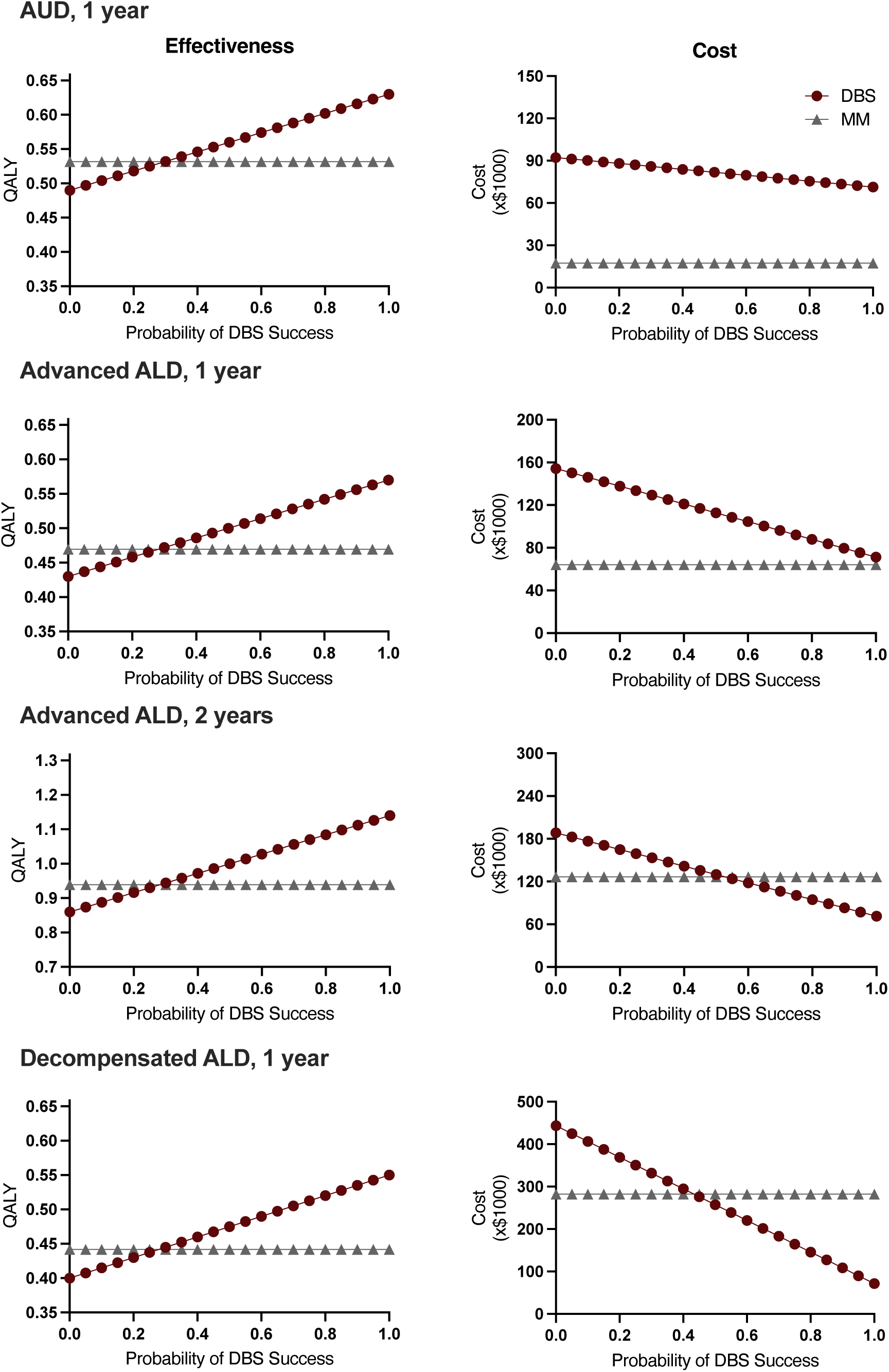
Threshold analysis of DBS probability of success compared to medical management (MM) for various patient populations. **Left,** effectiveness threshold analysis. **Right,** cost threshold analysis. In AUD patients, expected treatment cost plotted against DBS success probability shows that DBS remains significantly more expensive than MM over a one-year period. Expected QALYs plotted against DBS success probability show that MM and DBS result in equal QALYs when DBS is 29.8% successful. For patients with advanced ALD, DBS cost is close but remains higher than MM cost at a high DBS success probability over a one-year period. However, DBS becomes cost-saving over two-year periods when DBS success rate is > 53%. For patients with advanced decompensated ALD, MM and DBS are equally effective at a 27.9% DBS success rate but the cost of DBS becomes equivalent to MM at DBS success rate > 43.4%.

### Cost-effectiveness

Our probabilistic analysis combines the cost and effectiveness metrics to estimate the cost-effectiveness of DBS vs. MM for the different AUD populations. As expected, DBS is not cost-effective for AUD over one year at the 29.8% success rate, which corresponds to equivalent efficacy to MM. However, at success rates over 60%, DBS starts to become cost-effective but only at higher willingness-to-pay thresholds (Fig. 3A). For advanced ALD patients, DBS is cost-effective over one year at success rates ∼87% for $100,000/QALY willingness-to-pay threshold, and ∼60% success rate for $672,900/QALY willingness-to-pay threshold (Fig. 3B). For this same patient population, over 2 years, DBS is cost-effective at $100,000/QALY willingness-to-pay threshold when success rate is above 53% (Fig. 3C). For decompensated ALD patients, DBS dominates as the more cost-effective option over one year at success rates over 35% primarily due to its cost-saving properties independent of the willingness-to-pay threshold (Fig. 3D). It is worth mentioning that at low success rates (< 35%), DBS was never cost-effective regardless of willingness-to-pay threshold for any of the patient populations.

**Fig. 3.**
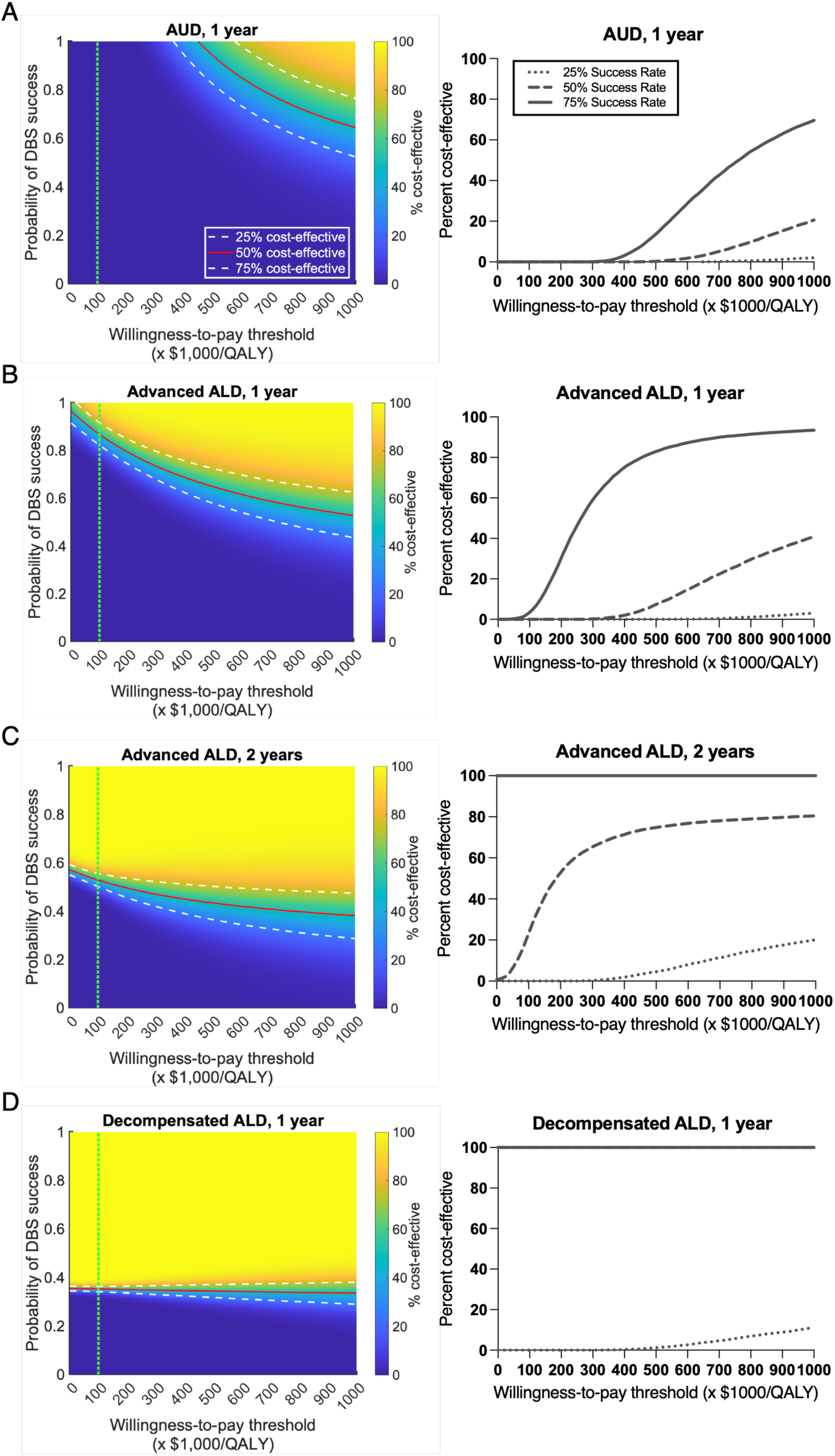
Probabilistic Analysis of DBS cost-effectiveness for various patient populations. **Left,** cost-effectiveness of DBS compared to MM at various willingness-to-pay thresholds and DBS success probability. **Right,** cost-effectiveness acceptability curve at 25%, 50%, and 75% DBS success probabilities. **A.** For AUD patients. DBS is not cost-effective at the $100,000/QALY willingness-to-pay threshold regardless of DBS’s probability of success. **B.** For advanced ALD patients over a one-year timeframe, DBS is only cost-effective at $100,000/QALY at success rates > 87%. **C.** For advanced ALD patients over a two-year timeframe, DBS is cost-effective at success rates > 53% at a $100,000/QALY threshold. **D.** For advanced decompensated ALD patients. DBS is cost-effective at >35% probability of success at a $100,000/QALY. The vertical green dashed line marks a willingness-to-pay of 100,000/QALY.

We also conducted similar analyses on AUD patients and any ALD (including early ALD with fibrosis stages 1 and 2, but the results were similar to the general AUD cost-effectiveness analysis due to the high prevalence and low costs associated with early ALD (data not shown).

Briefly, our analysis of the ICERs shows that at a $100,000/QALY willingness-to-pay threshold, DBS is not cost-effective for AUD patients without liver disease. For advanced ALD patients, DBS is cost-effective over two years at > 53% success rate. For decompensated ALD patients, DBS is cost-effective at > 35% success rate.

### Risk Analysis

The above analyses were conducted at standard DBS risks estimated in the movement disorders patient population. However, studies in AUD patients suggest that excessive alcohol use and/or ALD increase the risk of perioperative bleeding and other complications. Thus, we carried out a risk-based analysis of DBS cost-effectiveness in the advanced ALD and decompensated ALD groups (Fig. 4) [36, 37]. With DBS being cost-effective at 53% at standard DBS risk rate for the advanced ALD patient population, increased DBS risk starts to significantly impact cost-effectiveness when the risk rate exceeds by 4 folds compared to standard DBS risk. If DBS risk increases more than 16 times, MM dominates as more cost-effective at all success rates Fig. 4A). For decompensated ALD patients, the increased risk only begins to significantly impact DBS cost-effectiveness at risk 8 times greater than baseline. If these risks were increased by 32 folds, DBS still emerges as cost effective although at a higher probability of success (>50% vs. 35-40% success rate at lower risks).

**Fig. 4.**
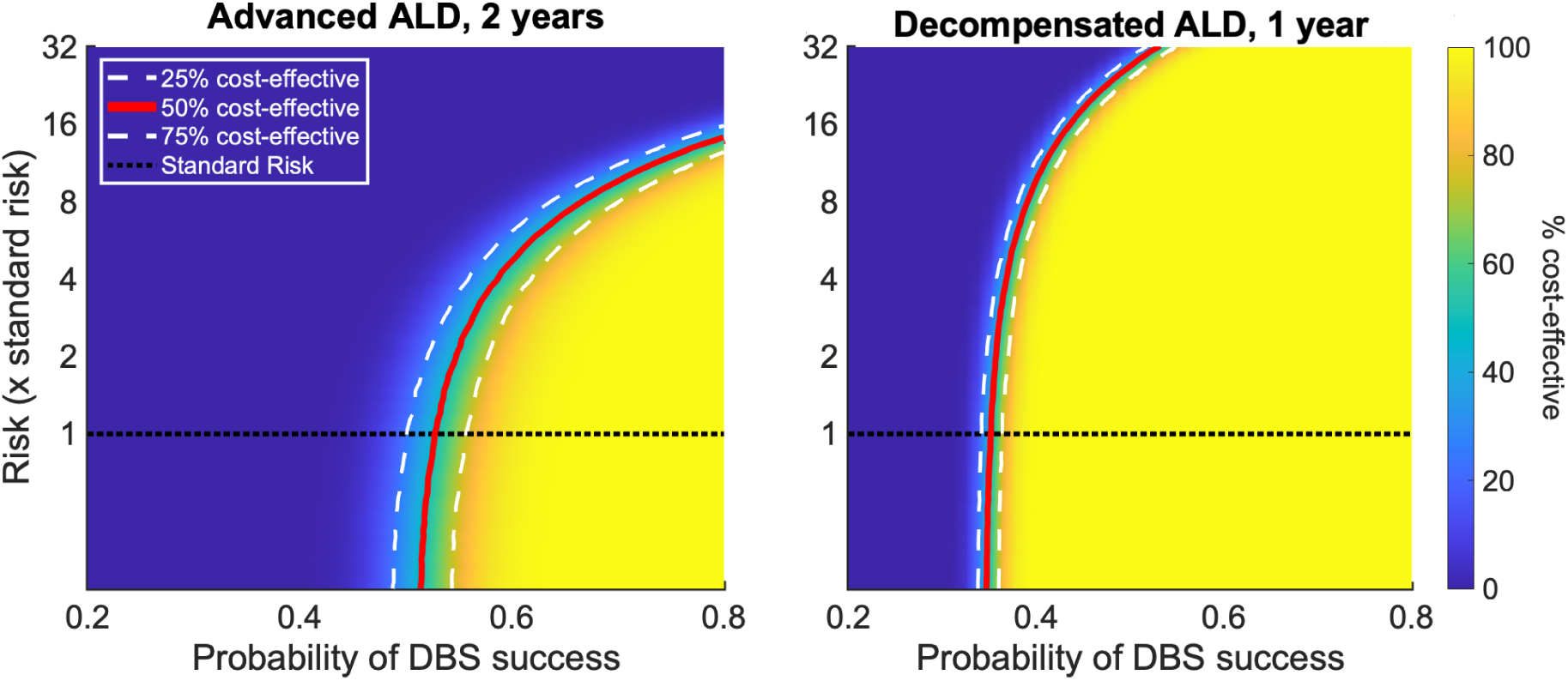
Cost-effectiveness of DBS compared to MM at various DBS-associated risks for advanced ALD (**A**) and advanced decompensated ALD (**B**). The red solid line indicates 50% cost-effectiveness, while the white dashed lines indicate 25% and 75% cost-effectiveness. The dotted green line represents standard DBS risk from movement disorder studies which is estimated to be ∼2.63%.

## Discussion

In this study, we utilized a probabilistic model to predict the cost-effectiveness of DBS as a treatment for AUD and ALD and explored how the increased risk associated with the procedure can affect its cost-effectiveness. We found that over a one-year time horizon, DBS is not cost-effective for AUD patients without ALD (at any success rate) or AUD patients with advanced ALD (fibrosis stage ≥ 3) when the DBS success rate is < 87%. However, DBS becomes cost-effective for patients with AUD and advanced ALD over 2 years at a success rate of > 55%. This is because QoL continues to increase, and societal costs decrease over the extended time period as the DBS cost is mostly limited to the first month of the intervention. Added long-term costs for DBS, such as battery replacement, usually occur over 3-5 years after initial DBS implantation. For patients with AUD and decompensated ALD, DBS is cost-effective as long as its success rate exceeds 35%.

A previous cost-effectiveness analysis for cocaine use disorder has also shown that the front-loaded costs of DBS hinder its cost-effectiveness in the short run [42]. In this study, DBS was shown to not be cost-effective at higher success probabilities over one year; however, over five years, it was shown to be significantly more cost-effective. A different DBS cost-effectiveness analysis for opioid use disorder suggested that DBS would be cost-effective if its success rate is greater than 49% [41]. This is consistent with our data especially given that advanced liver disease is associated with high health care and social costs from decompensated cirrhosis (infections, bleeding, ascites, encephalopathy), liver transplant, and hepatocellular carcinoma.

Peri-operative risk is increased in patients with AUD, including advanced ALD patients [36, 48, 49]. Studies show that patients with either liver disease or a history of heavy alcohol use are at an increased risk of bleeding and other complications. Thus, it is likely that DBS surgery in AUD patients, especially those with ALD, will be associated with increased risks. Nonetheless, our analysis shows that DBS remains cost-effective with up to 18-fold increase in DBS-associated risk, as long as DBS success rate is around or greater than 41%. Besides cost-effectiveness, and despite the increased surgical risks, the absence of alternative treatments for AUD warrants investigating DBS as a treatment option for patients with AUD and ALD given the poor prognosis when AUD is left untreated [28, 50].

As aforementioned, DBS studies for substance use disorders including AUD are still preliminary but hint to potential efficacy [27, 51, 52]. In a case series of 5 patients with severe AUD implanted with VS DBS, 2 patients showed complete remission for several years, 1 patient abstained for 1.5 years then had several relapses, and 2 patients continued to relapse and died few years later, presumably from continued alcohol use [51]. In addition, a recent trial of ventral striatum DBS in 6 patients with AUD showed ∼74% reduction in alcohol consumption[52]. Finally, another recent randomized clinical trial, though underpowered for the primary outcome, showed increased number of abstinent days and reduced alcohol consumption in the DBS active group [27].

The current cost-effectiveness analysis for DBS in AUD patients has several limitations. We used parameter estimates from multiple independent clinical trials and, as a result, included distinct study populations selected with different eligibility criteria, potentially introducing confounding population heterogeneity. Thus, conservative ranges were utilized in all analyses to ensure robustness of the results. Given the limited literature on the treatment of alcohol use disorder in patients with ALD, we used the same MM success rate and increase in QoL for both AUD and ALD populations as reported by previous studies [35, 39]. We also ensured that the QoL estimates we used were from studies that applied consistent methodologies within the different patient populations. Another limitation is that the success or failure of DBS was computed as a binary measure. For example, our model does not consider DBS success as a continuous measure with continuous effects on QoL. Finally, our analysis assumes that the QoL of patients after successful treatment is consistent between medical management and DBS independent of complication risks.

As DBS is not well studied for the treatment of SUDs, costs and complications were adopted from trials that used DBS for the treatment of movement disorders. DBS costs for patients are likely lower than we report as these are mitigated by insurance packages. A one-year time horizon does not fully account for all relevant costs and effects of DBS and MM. While most of the cost associated with DBS is usually upfront, at the time of DBS implantation, DBS is usually associated with additional costs related to battery replacement 3-5 years after initial system implantation, and regular follow-up visits. However, these additional costs are likely to be mitigated by efficacy should DBS proves effective.

This analysis suggests that if efficacious, DBS may be cost-effective for selected subpopulations of patients with AUD. It also supports the need for further clinical trials focused on the safety and efficacy of DBS for AUD and ALD patients.

## Data Availability

All data produced in the present study are available upon reasonable request to the authors

## Abbreviations

DBS: Deep Brain Stimulation
AUD: alcohol use disorder
ALD: alcoholic liver disease
MM: medical management
QoL: quality of life
QALY: quality-adjusted life year

## Acknowledgments

We would like to thank Dr. Jovan Julien for providing some data on the ALD-associated costs.

## Author Contributions

Conceptualization: OA, HJ, KM

Formal analysis: OA, CL

Funding acquisition: HJ, KM

Investigation: OA, KM

Methodology: OA, CL, HJ, KM

Software: OA, CL

Supervision: HJ, KM

Visualization: OA, CL, HJ, KM

Writing -original draft: OA, KM

Writing - review & editing: OA, CL, HJ, KM

